# Identification and Evaluation of Serum Protein Biomarkers Which Differentiate Psoriatic from Rheumatoid Arthritis

**DOI:** 10.1101/2020.06.23.20138552

**Authors:** Angela Mc Ardle, Anna Kwasnik, Agnes Szenpetery, Melissa Jones, Belinda Hernandez, Micheal Meenagh, Andrew Parnell, Wilco de Jager, Sytze de Roock, Oliver FitzGerald, Stephen Pennington

## Abstract

**Objectives:** To identify serum protein biomarkers which might separate early inflammatory arthritis (EIA) patients with psoriatic arthritis (PsA) from those with rheumatoid arthritis (RA) to provide an accurate diagnosis and support appropriate early intervention.

**Methods:** In an initial protein discovery phase, the serum proteome of a cohort of patients with PsA and RA was interrogated using unbiased liquid chromatography mass spectrometry (LC-MS/MS) (n=64 patients), a multiplexed antibody assay (Luminex) for 48 proteins (n=64 patients) and an aptamer-based assay (SOMAscan) targeting 1,129 proteins (n=36 patients). Subsequently, analytically validated targeted multiple reaction monitoring (MRM) assays were developed to further evaluate those proteins identified as discriminatory during the discovery. During an initial verification phase, MRM assays were developed to a panel of 150 proteins (by measuring a total of 233 peptides) and used to re-evaluate the discovery cohort (n=60). During a second verification phase, the panel of proteins was expanded to include an additional 23 proteins identified in other proteomic discovery analyses of arthritis patients. The expanded panel was evaluated using a second, independent cohort of PsA and RA patients (n=167).

**Results:** Multivariate analysis of the protein discovery data revealed that it was possible to discriminate PsA from RA patients with an area under the curve (AUC) of 0.94 for nLC-MS/MS, 0.69 for Luminex based measurements; 0.73 for SOMAscan analysis. During the initial verification phase, random forest models confirmed that proteins measured by MRM could differentiate PsA and RA patients with an AUC of 0.79 and during the second phase of verification the expanded panel could segregate the two disease groups with an AUC of 0.85.

**Conclusion:** We report a serum protein biomarker panel which can separate EIA patients with PsA from those with RA. We suggest that the routine use of such a panel in EIA patients will improve clinical decision making and with continued evaluation and refinement using additional patient cohorts will support the development of a diagnostic test for patients with PsA.

---

Psoriatic Arthritis (PsA) is a form of inflammatory arthritis (IA) affecting approximately 0.25% of the population [1-4]. It is a highly heterogeneous disorder associated with joint damage, disability, disfiguring skin disease and poor patient-related quality of life outcome measures [4]. Inherently irreversible and frequently progressive, the process of joint damage begins at, or before, the clinical onset of disease. Indeed, structural joint damage, which is likely to result in joint deformity and disability, is present in 47% of patients within 2 years of disease onset [3, 5]. Reductions in quality of life and physical function are comparable to rheumatoid arthritis (RA) and compounded by the presence of chronic disfiguring skin disease [6-9]. Direct and indirect health costs pose a significant economic burden on society and rise with severe physical dysfunction [9].

Early diagnosis and management of PsA leads to better long-term outcomes however with no diagnostic laboratory test available, the diagnosis is often delayed or missed and this has significant consequences for individuals with PsA [10-12]. At disease onset PsA often resembles other forms of arthritis including RA. Despite the clinical similarities between PsA and RA, their distinctive pathologies often require different treatments. For example, drugs targeting the IL-12/IL-23 and IL-17 pathway which are highly effective in psoriasis and PsA, are ineffective in RA, while drugs targeting B cells such as rituximab are effective in RA but have not been proven beneficial in PsA [4, 13].

PsA is most often diagnosed when a patient presents with musculoskeletal inflammation in the presence of psoriasis and in the absence of rheumatoid factor (RF). However, a clear diagnosis can be difficult as up to 10% of PsA patients may have RF or Anti-Citrullinated Peptide Antibody (ACPA) and joint involvement may precede the development of skin or nail psoriasis in 15 % of patients with PsA [14]. The “<u>C</u>l<u>AS</u>sification criteria for <u>P</u>soriatic <u>AR</u>thritis” (CASPAR) are accepted as having high sensitivity (98.7 %) and specificity (91.4 %) in classifying patients with long-standing PsA. CASPAR shows reduced sensitivity in patients with early disease (87.4 %) though specificity is improved (99.1 %) [15]. CASPAR are valid when including patients in research studies or in clinical trials, but it is recognised that they should not be used for diagnosis and are of little value therefore in a primary care or dermatology setting where specialist rheumatological expertise is very often not readily available [4, 16]. An effective clinical laboratory test is needed to improve diagnosis and clinical decision making in PsA.

Ideally, a clinical laboratory test should be based on an easily accessible biological sample such as blood [10]. Therefore, we set out to discover serum-based biomarkers that could discriminate between patients with PsA or RA. With advances in multiplexed technologies, it has become possible to simultaneously measure multiple analytes. However, in complex bio-fluids such as serum, it is apparent that no single technological platform is capable of measuring the entire protein content of a given sample [3, 4, 17]. For this reason, we undertook a comprehensive and complementary analysis of the serum proteome in an EIA cohort. We used unbiased nLC-MS/MS to identify the more abundant, differentially expressed proteins. In parallel, the SOMAscan and Luminex assays were employed to target low abundant proteins not easily detectable by nLC-MS/MS. Statistical analysis revealed that proteins identified by nLC-MS/MS were the most useful in discriminating individuals with PsA from those with RA. Hence in subsequent steps we prioritised these proteins for further investigation. Using multiple reaction monitoring (MRM), a form of targeted mass spectrometry, we designed a two-phase verification study: in the first phase, we measured a panel of 150 proteins identified in the discovery cohort and, in the second phase, we measured an expanded panel of 173 proteins in an independent cohort. **Figure 1** provides an overview of the workflow of the study.

**Figure 1.**
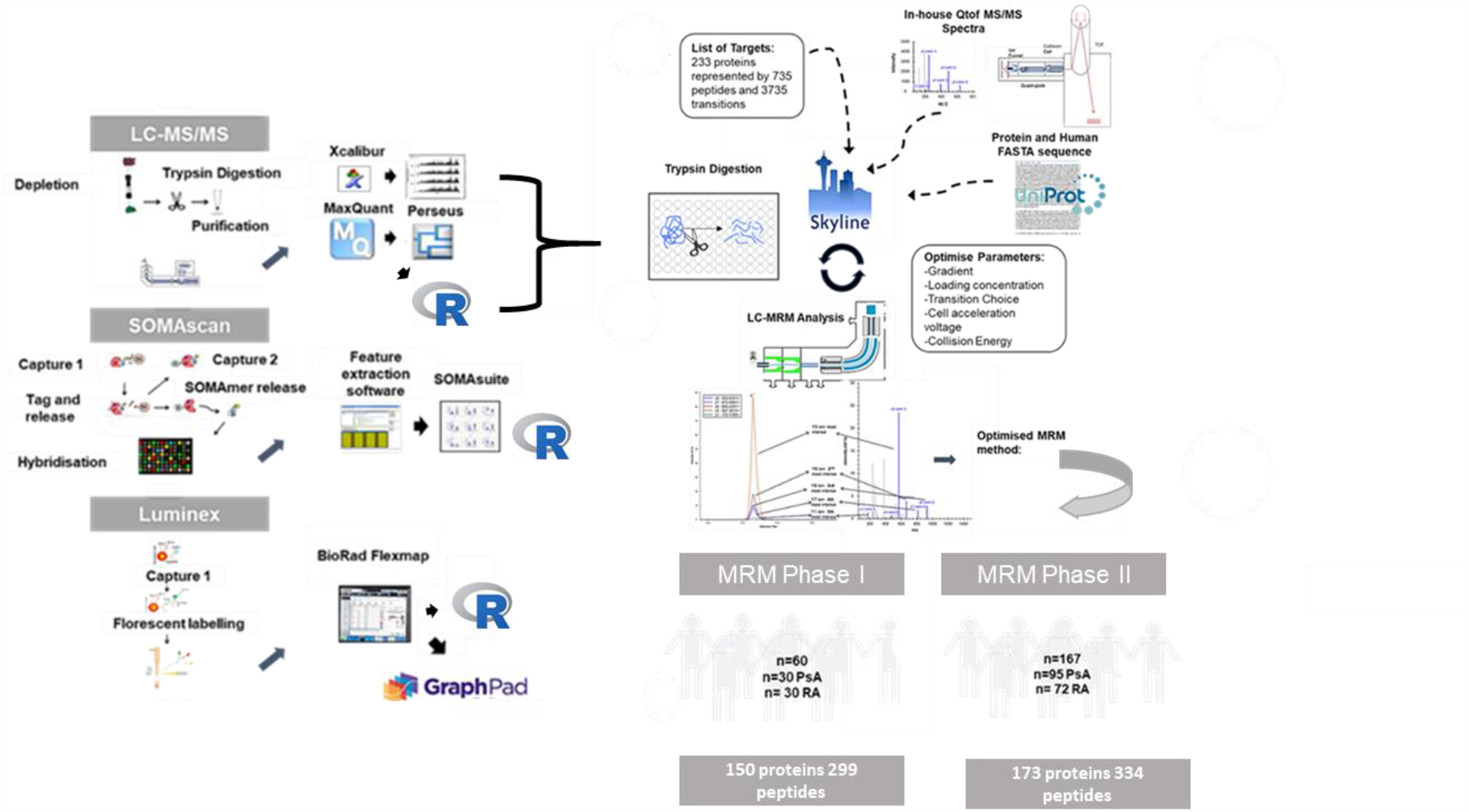
Overview of experimental worklfow. Three different platforms were employed: nLC-MS/MS, Luminex xMAP and SOMAscan for biomarker discovery. Resulting data was analysed by univariate and multivariate analysis. Details of the versions of software are described in methodology section. Putative biomarkers identified by nLC-MS/MS proteins were brought forward for MRM assay development which was divided into two phases. During phase I, it was possible to develop an assay to 150 proteins which were measured in the discovery cohort. During phase II, an assay was developed to 173 proteins which were measured in an independent verification cohort.

## Patients and Methods

### Patients

In the discovery phase and initial verification phase, a total of 64 patient samples were used and a full description of the extensive clinical characterisation of the cohort can be found in Szentpetery *et al*. [18]. In brief, recent-onset (symptom duration <12 months), treatment naïve PsA and RA patients with active joint inflammation, aged 18 to 80 years were enrolled. PsA patients (n=32) fulfilled the CASPAR criteria [19] and patients with RA (n=32) met the 2010 ACR/EULAR classification criteria [20]. Baseline serum samples were obtained from each patient using standard methodology, aliquoted and frozen at -70°C (**Suppl Doc 1**). The study was approved by St.Vincent’s Healthcare Group Ethics and Medical Research Committee and patients were enrolled only after agreeing to participate in the study and having given their informed consent.

Samples from a total of 167 patients were used in the second verification phase. There were 95 patients recruited from a cross-sectional cohort of established PsA patients, all meeting CASPAR criteria and 72 patients recruited from the RA Biologics Registry of Ireland (RABRI) who all met 2010 ACR/EULAR classification criteria and had similar levels of active disease, as the PsA patients. Again, baseline serum samples were obtained, aliquoted and frozen at - 70°C.

### Label Free nLC-MS/MS Analysis

A detailed description of the unbiased LC-MS/MS workflow can be found in *Mc Ardle et al*. [10] In brief serum samples (1,700 ug) were depleted of the 14 most abundant serum proteins (albumin, transferrin, haptoglobin, IgG, IgA, 1-antitrypsin, fibrinogen, β2-macroglobulin, 1-acid glycoprotein, complement C3, IgM, apolipoprotein AI, apolipoprotein AII, and transthyretin)) using the Agilent Multiple Affinity Removal System comprising a Hu-14 column (HuMARS14) (4.6 × 100 mm; Agilent Technologies, 5188-6557) on a Biocad Vision Workstation. Depleted fractions (containing 50 µg protein) were reduced, denatured and alkylated prior to trypsinization. The digested samples were desalted and purified using C18 resin pipette stage tips. Purified samples were dried under vacuum and resuspended in mass spectrometer compatible buffer A (3% ACN, 0.1% formic acid) [21, 22]. Label-free nano-flow LC-MS/MS analysis was performed on a Q-Exactive mass spectrometer equipped with a Dionex Ultimate 3000 (nano-RSLC) chromatography system (Thermo Fisher Scientific). Two microliters (equivalent to 2 µg of digested protein) of each sample was injected onto a fused silica emitter separated by an increasing acetonitrile gradient over 101.5 min (flow rate of 250 nL/min) [10].

### Bioinformatic Data Analysis

As previously reported nLC-MS/MS data were visually inspected using XCalibur software (2.2 SP1.48). MaxQuant (1.4.12) was then used for quantitative analysis of the LC-MS/MS data (Thermo Scientific. Raw) while Perseus software (1.5.0.9) supported statistical analysis [10, 23].

### SOMAscan Analysis

Individual patient serum samples were subjected to a multiplexed aptamer-based assay (SOMAscan) developed by *Gold et al*. to measure the levels of 1129 proteins as previously reported [10].

### Luminex Analysis

Individual serum samples were subjected to an in-house developed and validated multiplexed immunoassays measuring 48 analytes with Luminex xMAP proteomics technology (Austin TX, USA). The assays, including the analyses, were undertaken as previously described by the Multiplex Core Facility Laboratory of Translational Immunology LTI, in the University Medical Centre Utrecht [10].

### MRM design and Optimisation

The development and optimisation of MRM assays was performed using Skyline software (version 3.6.0.1062) (MacCoss laboratory, Washington DC) [24]. Assays were developed to proteotypic peptides for all proteins of interest where peptides showed no missed cleavages or ‘ragged ends’ and sequence length was between 7-25 amino acids. Where possible, peptides sequences with reactive cysteine (C) or methionine (M) residues were avoided but not excluded. An MRM assay was deemed to be analytically validated when it demonstrated the following characteristics: dot product ≥0.8; signal to noise ≥10; data points under the curve ≥10 [25]; and percentage coefficient of variance showing a retention time ≤ 1 % and area ≤ 20 % [26]. The majority of MRM assays developed significantly exceeded these criteria.

### Sample Preparation for LC-MRM Analysis

#### Initial Phase

Crude serum (2 µl) was added to the wells of 96 deep well plates (Thermo) and diluted 1 in 50 with NH_4_CO_3_ (Sigma). Rapigest denaturant (Waters), resuspended in 50mM NH_4_CO_3_ to give a stock solution of 0.1% w/v 50 ul stock solution, was added to each sample so that the final concentration of Rapigest was 0.05 %. Plates were covered with adhesive foil (Thermo) and samples were incubated in the dark at 80 °C for 10 minutes (min). After incubation plates were centrifuged at 2,000 relative centrifugal force (rcf) at 4 °C for 2 min to condense droplets. Subsequently, dithiothreitol (DTT) was added to each sample at a final concentration of 20 mM. Samples were then incubated at 60°C for 1 hr followed by centrifugation at 2,000 rcf at 4 °C for 2 min. Next iodoacetamide (IAA) was added to each sample to give a final concentration of 10 mM and plates were incubated at 37 °C in the dark for 30 min. Plates were again centrifuged at 2,000 rcf at 4 °C for 2 min and samples were then diluted with LC-MS/MS grade H_2_O to give a final concentration of 25 mM NH_4_CO_3_. Trypsin (Promega) was added to each sample so that the protein enzyme ratio was 25:1. The reaction was stopped with the addition of 2 ul of neat trifluoroacetic acid (TFA, Sigma) to each sample and incubated for a further 30 min at 37 °C. In order to pellet Rapigest, digests were transferred from 96 well plates to 1.5 ml low-bind Eppendorf tubes and centrifuged for 30 min at 12,000 rcf. Supernatants were removed and transferred into clean Eppendorf tubes and lyophilised by speed vacuum at 30 °C for 2 hr. Lyophilised samples were stored at -80 °C until further use. For the ***second phase:*** the denaturant used previously (rapigest) was substituted with 25 µL denaturant solution comprising 50 % trifluoroethanol (TFE) in 50 mM NH_4_HCO_3_with 10 mM DTT and this mitigated the need for the high-speed spin and transfer of supernatant which represented an additional processing step less compatible with 96 well plate workflows.

#### LC-MRM Analysis

MRM analysis was performed using an Agilent 6495 triple quadrupole (QqQ) mass spectrometer with a JetStream electrospray source (Agilent) coupled to a 1290 Quaternary Pump HPLC system. Peptides were separated using analytical Zorbax Eclipse plus C18, rapid resolution HT: 2.1 x 50 mm, 1.8um, 600 Bar columns (Agilent) before introduction to the QqQ. A linear gradient of acetonitrile (99.9 % ACN & 0.1 % FA) 3 -75 % over 17 mins was applied at a flow rate of 0.400 µl/min with a column oven temperature of 50 °C. Source parameters were as follows; gas temp: 150 °C, gas flow 15 l/min, nebuliser psi 30, sheath gas temp 200 °C and sheath gas flow 11 l/min. Peptide retention times and optimised collision energies were supplied to MassHunter (B0.08 Agilent Technologies) to establish a dynamic MRM scheduled method based on input parameters of 80 millisecond (ms) cycle times and 2 min retention time windows. The percentage coefficient of variance (% Cv) of biological and technical replicates was used as a measure of variance and was calculated using the following standard calculation: % CV = (standard deviation/mean) x 100.

#### Enzyme linked Immunosorbent Assay Analysis

CRP levels were evaluated at St Vincent’s University Hospital, Dublin using an automated CRPL3 Tina-quant assay (Roche Diagnostics, GmbH).

#### Statistical Analysis

Graphpad Prism software package (7.00) was used to investigate the statistical significance of Luminex data whereas SOMAsuite (1.0) was used to analyse SOMAscan data. The ability of quantified proteins/peptides to predict the diagnosis (PsA or RA) of individual patients was assessed using the Random forest package in R (version 3.3.2). The most important variables in providing the area under the receiver operating curve were selected by using the variable importance index and the Gini decrease in impurity was used to assess the importance of each variable. All area under the curve (AUC) values were obtained using the ROCR package in R (version 3.3.2).

## Results

### Patient sample characterisation and study design

For the discovery of candidate novel protein biomarkers, serum samples were collected at baseline from early onset, treatment naïve PsA (n=32) and RA (n=32) patients. Samples from a second independent cohort (PsA n=95; RA n=72) were used to confirm the performance of the putative markers identified during discovery. While these PsA and RA patients may have been on treatment at time of baseline serum sampling, there were similar levels of active disease (as reflected by CRP, ESR and joint counts) in both patient groups. Key demographic and clinical features of all patients are summarised in **Table 1**.

**Table 1.**
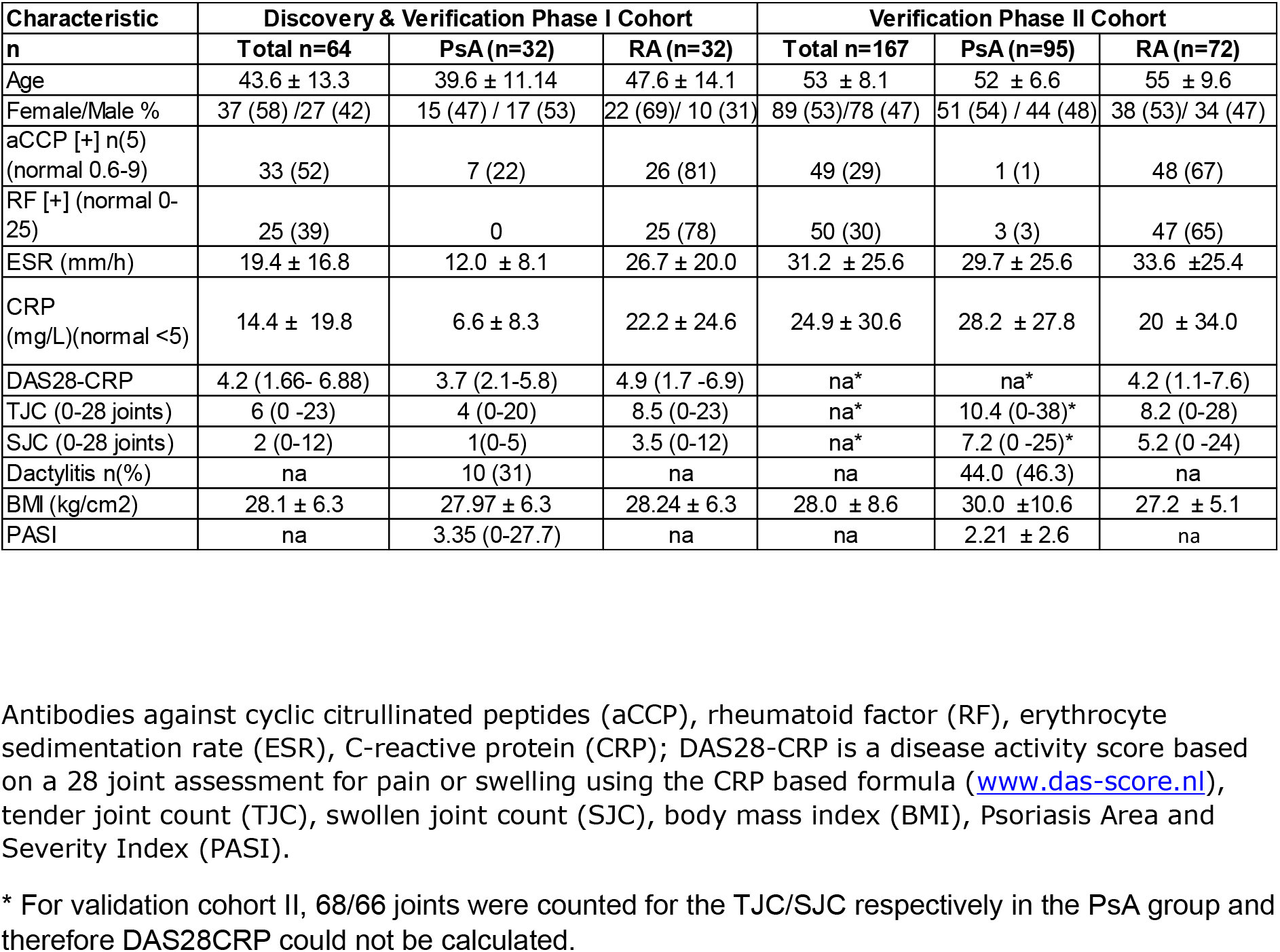
Baseline demographic and clinical parameters of the discovery and verification cohorts.

### Unbiased nLC-MS/MS based protein analysis

To investigate differential serum protein expression between patients with PsA and RA, individual serum samples which had been depleted of high abundance serum proteins were analysed by nLC-MS/MS using a QExactive Hybrid Quadrupole-Orbitrap Mass Spectrometer mass spectrometer. A total of 451 proteins were identified of which 121 were identified in all 64 individual serum samples. Univariate analysis was applied to the 121 commonly identified proteins and multivariate analysis was applied to the complete data set. Univariate analysis (Student t-test using a Benjamini-Hochberg FDR of 0.01) showed that 66 proteins were significantly differentially expressed between PsA and RA (**Suppl. Table 1)**. Unsupervised hierarchical cluster and principle component analysis on these 66 proteins revealed the overall differences/similarities between serum protein levels in the individual PsA and RA patients; clear within group clustering and between group separations could be observed (**Figure 2**). Random forest analysis of data from 451 proteins identified in the 64 patient samples demonstrated that patients with PsA and RA could be differentiated with an AUC of 0.94 (**Table 2**) (ROC plot **Suppl. Fig 1A**). Together these data strongly suggest that there is a difference in the serum protein profiles between newly diagnosed PsA and RA patients. The top 50 proteins providing the AUC are listed in (**Suppl. Table 2)**.

**Figure 2.**
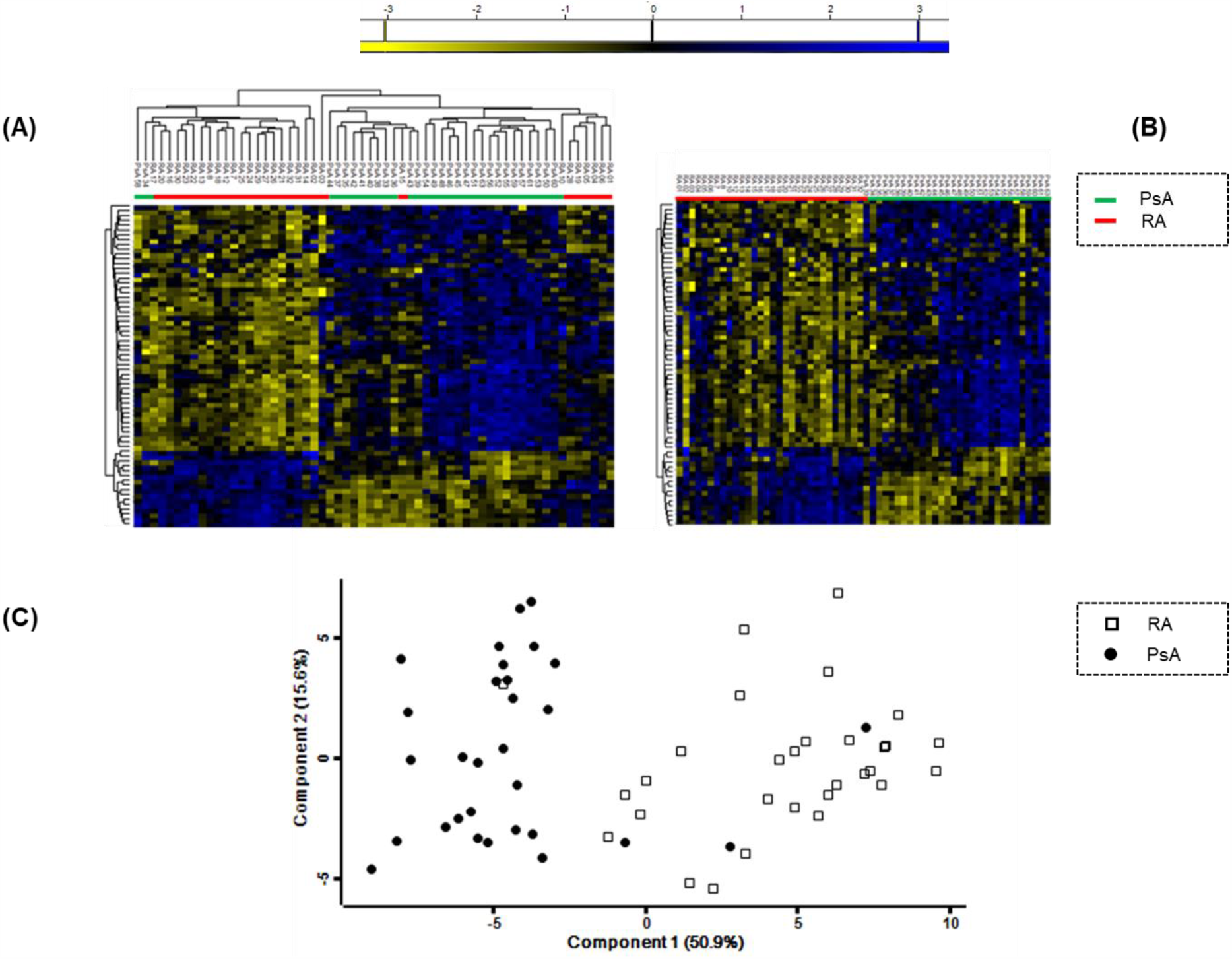
Association of protein signatures with diagnosis as shown by (**A**) unsupervised hierarchical cluster analysis (HCA), (**B**) supervised HCA and (**C**) Principal component analysis Plots were generated on differentially expressed proteins between PsA (n=30) and RA (n=30) patients (p≤ 0.01, Benjamin Hochberg FDR).

**Table 2.** Determination of protein signatures to predict diagnosis in patients with early PsA and RA. Area under the curve (AUC) values were generated using the predicted probabilities from the random forest model used to discriminate between the groups

| **Platform** | **n** | **Correctly Predicted** | **AUC** |
| --- | --- | --- | --- |
| LC-MS/MS | 60 | 55/60 | 0.94 |
| SOMAscan | 36 | 26/36 | 0.73 |
| Luminex | 64 | 43/64 | 0.69 |

### SOMAscan and Luminex targeted protein analysis

To extend the breadth and depth of proteome coverage afforded by nLC-MS/MS, serum samples were subjected to analysis on 2 complementary protein measurement platforms. SOMAscan analysis supported the quantification of 1129 proteins in a subset of the patient samples PsA (n=18) and RA (n=18). Univariate analysis revealed that 175 proteins were significantly differentially expressed between PsA and RA patients (**Suppl. Table 3**). Multivariate analysis of the data obtained from the SOMAscan analysis revealed that it was possible to discriminate PsA from RA patients with an AUC of 0.73 (**Table 2**) (ROC plot **Suppl. Fig 1B**).

Based largely on their known importance in PsA and RA (from a literature review [3]), 48 proteins were selected for analysis using in-house developed multiplexed Luminex assays [10]. Of the 48 proteins targeted, 23 were identified in every sample. T-tests revealed that 4 proteins; IL-18 (p ≤ 0.001), Il-18 binding protein (BPa) (p ≤ 0.05), hepatocyte growth factor (HGF) (p ≤ 0.05) and tumour necrosis factor receptor superfamily member 6 (FAS) (p ≤ 0.05) were differentially expressed between PsA and RA samples (**Suppl Fig 2**). Random forest analysis of the Luminex data showed that patients could be segregated with an AUC of 0.64 (**Table 2**) (**Suppl Fig 1C**). In comparison to the nLC-MS/MS analysis, the candidate protein biomarker discovery by both SOMAscan and by Luminex yielded data sets with reduced predictive power and therefore the subsequent evaluation process was streamlined to focus only on proteins identified by nLC-MS/MS.

### LC-MRM verification of nLC-MS/MS identified biomarkers

Multiple reaction monitoring (MRM) is a targeted MS technology which is increasingly used to support candidate biomarker evaluation following LC-MS/MS and other protein discovery approaches. Both the cost of MRM analysis and the time required to develop and optimise MRM assays are considerably less than antibody-based methods [27]. For these and other reasons, MRM based measurement of the nLC-MS/MS-identified proteins represented an attractive approach for verification and evaluation of their biomarker performance. The multiplexing capabilities afforded by MRM facilitated the development of an assay that included the top-ranking discriminatory candidate proteins from univariate and multivariate analysis of the nLC-MS/MS discovery data described above but also allowed for the inclusion of additional proteins identified previously during studies of pooled patient samples (data not shown). During a first verification phase, a total of 233 proteins represented by 735 peptides and 3735 transitions (5 per peptide) were brought forward for MRM assay development. Of the 233 proteins brought forward it was possible to develop analytically validated assays for 150 of them, represented by 299 peptides; the remaining candidates were either undetectable in crude serum or had assays that did not meet our analytical criteria plasma (Of the 50 priority proteins listed in Suppl Table 2, 33 were included in the assay). This MRM assay panel was then used to verify the candidate proteins in the discovery cohort (n=60). It is noteworthy that to minimise any technical bias both the pre-analytical processing and MRM analysis of was undertaken in a randomised manner. Random forest analysis revealed that using this MRM assay panel, it was possible to discriminate PsA from RA with an AUC of 0.79 (**Figure 3A**). During a second verification phase, an additional 23 proteins, identified as being discriminatory in other analyses of inflammatory arthritis patients, were added to the initial MRM assay panel, giving a new total number of proteins of 173 (represented by 334 peptides) [28]. This expanded panel was used to measure protein candidates in an independent verification cohort of 95 PsA and 72 RA patients (**Table 1**). Seven synthetic isotopically labelled (SIL) peptides were incorporated into the assay to control for potential analytical variation. Summed intensity values from the SIL peptides were used to normalise patient data. Random forest analysis revealed that PsA patients could be separated from those with RA with an AUC of 0.85 (**Figure 3B**). The proteins ranked most important in providing the AUC values are reported in **Table 3**. The data demonstrate clear overlap between proteins used to segregate PsA and RA patients included in the discovery and verification cohorts. The differential expression levels of these overlapping proteins are illustrated in **Suppl Fig 3**. To this end, Alpha-2-HS-glycoprotein **(A2AGL)**, Alpha-1-antichymotrypsin **AACT)**, Haptoglobin **(HPT)**, Haptoglobin-related protein **(HPTR)**, Rheumatoid factor C 6 light Chain **(V-Kappa-1)** were found to be significantly upregulated in RA patients compared to PsA when measured by MRM in the cohorts included in phase I and phase II. Alpha-1-acid glycoprotein **(A1AG)** and coagulation factor XI **(FA11)** were also found to be upregulated in RA compared to PsA during both rounds of verification, however the observation only reached significance during the second phase. In contrast thrombospondin 1 (**TSP1**) was found to slightly upregulated in RA patients during the first phase but significantly upregulated in PsA patients during the second evaluation phase. These observations confirmed the validity and performance of the initially developed classifier, but also demonstrated how further development of the assay enhanced performance of the predictive algorithm. This on-going evolution of the MRM assay panel and associated artificial intelligence (AI) and machine learning algorithms represents a new and powerful approach to biomarker development.

**Figure 3.**
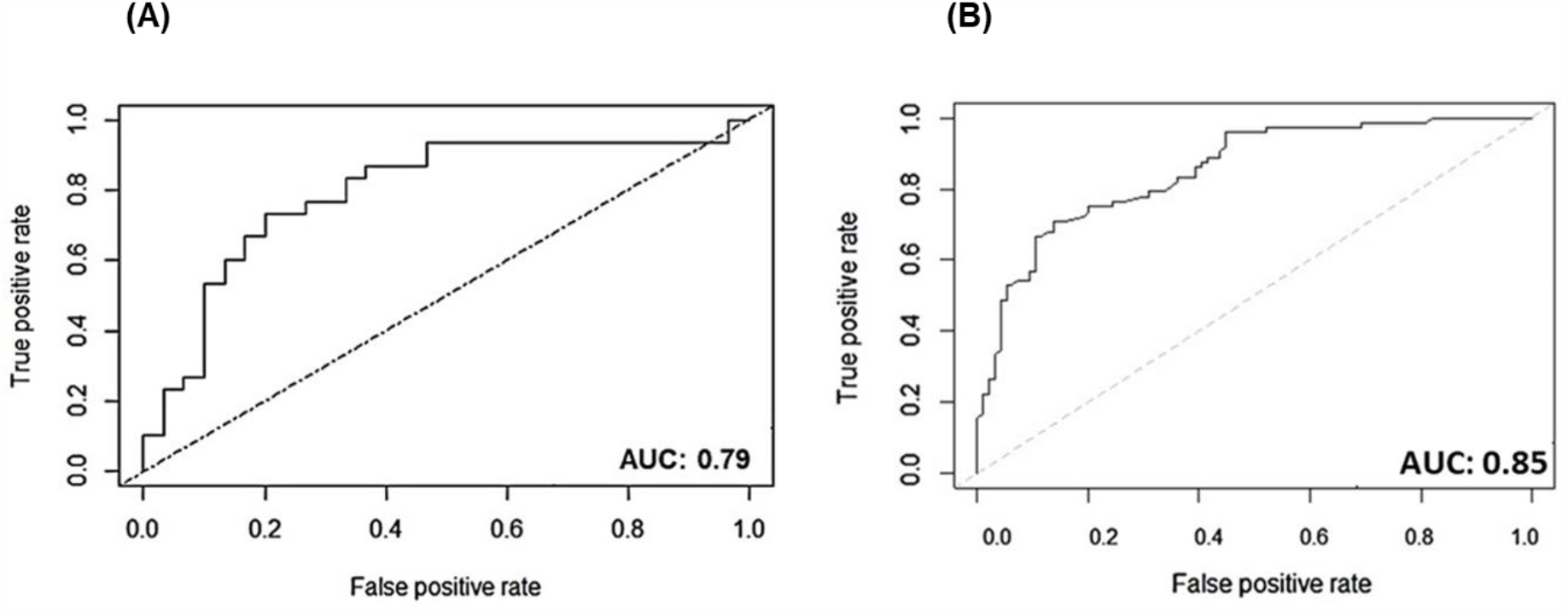
Reciever operating characterstics (ROC) curve for (A) performance of protein signatures in discovery cohort (MRM verifcaiton phase I, Total=60: 30 PsA; 30 RA) and (B) in an independent verfication cohort (MRM verificaiton phase II, Total=167: 95 PsA; 72 RA).

**Table 3.**
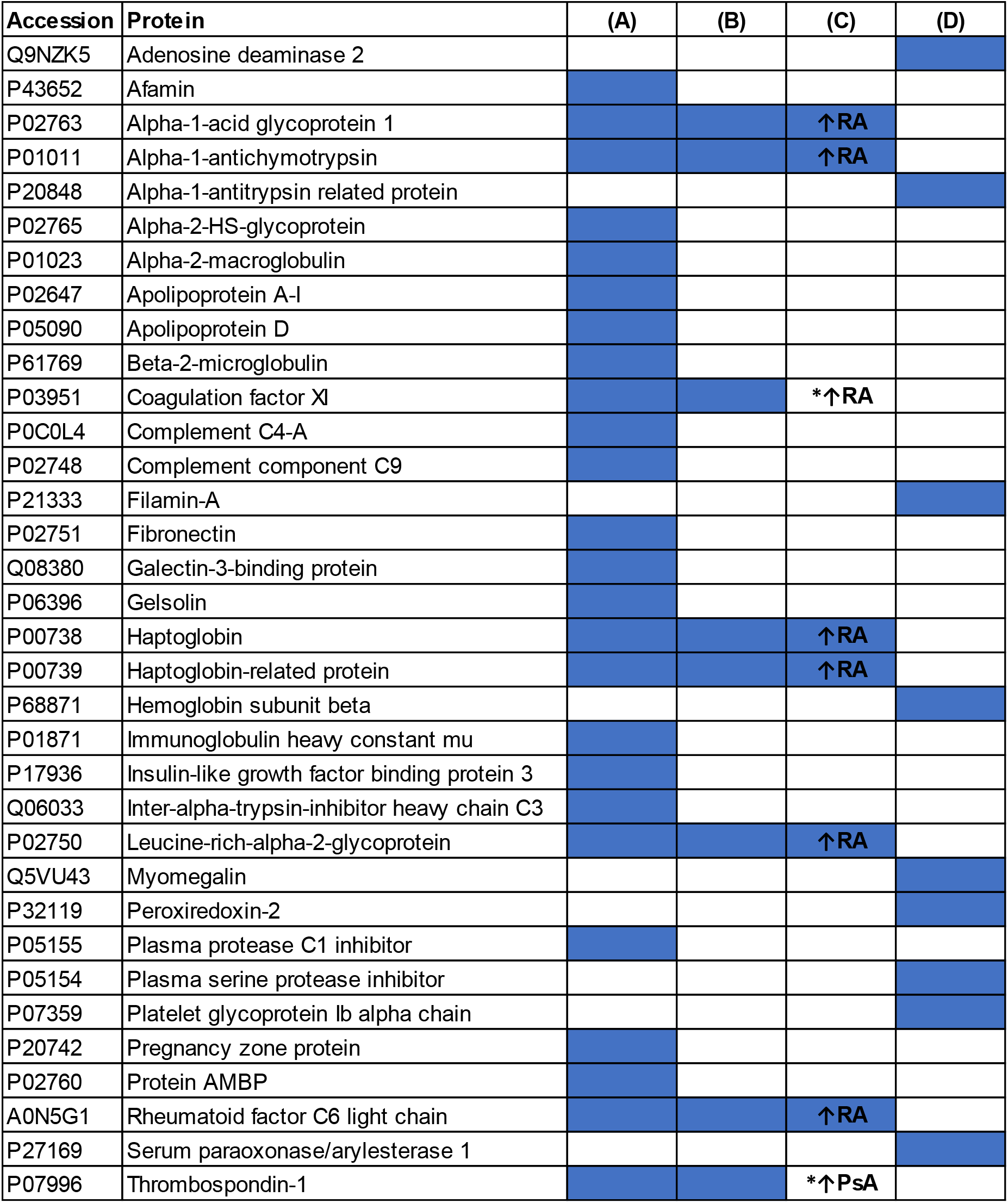
The top 34 proteins providing the AUC of 0.85. Proteins listed alphabetically were rerpresented by proteotypic peptides when measured by MRM **(A)** Of the 34 listed, 25 were measured in both MRM phase I and phase II assays. **(B)** Of those 25 proteins, 8 proteins were top contributers to the AUC in generated during phase I (AUC 0.79) and phase II (AUC 0.85). **(C)** Of the 8 protiens in (B), 6 were significantly urpegualted in RA (un-pairded T test, see **Suppl Fig 4**). *Coagulation factor X1 and thrombospndin did not reach statstical signficance during phase I analysis (**Suppl Fig 4)** Of the 34 proteins listed in **(D)**, 9 proteins providing AUC 0.85 were exclusively present in the MRM phase II assay.

Finally, there are at least two potential routes to implementing a multiplexed protein biomarker panel in the clinical setting. One is to use MRM assays and the other to develop antibody-based assays to the proteins of interest. To explore the extent to which MRM data may align with ELISA we compared our MRM data for CRP with results obtained by standard clinical laboratory ELISA. MRM measurements were compared to the ELISA measurements in the 60 samples from the discovery set. It was not surprising to find that serum levels of CRP were significantly upregulated in patients with RA as compared to those PsA when measured by both ELISA (p ≤ 0.0009) and MRM (p ≤ 0.0006) (**Figure 4B)**. Furthermore, CRP values from both platforms were strongly correlated (R^2^ = 0.8345) (**Figure 4C)** indicating that a measurement made by MRM can give a value similar to that obtained by an existing immuno-assay.

**Figure 4.**
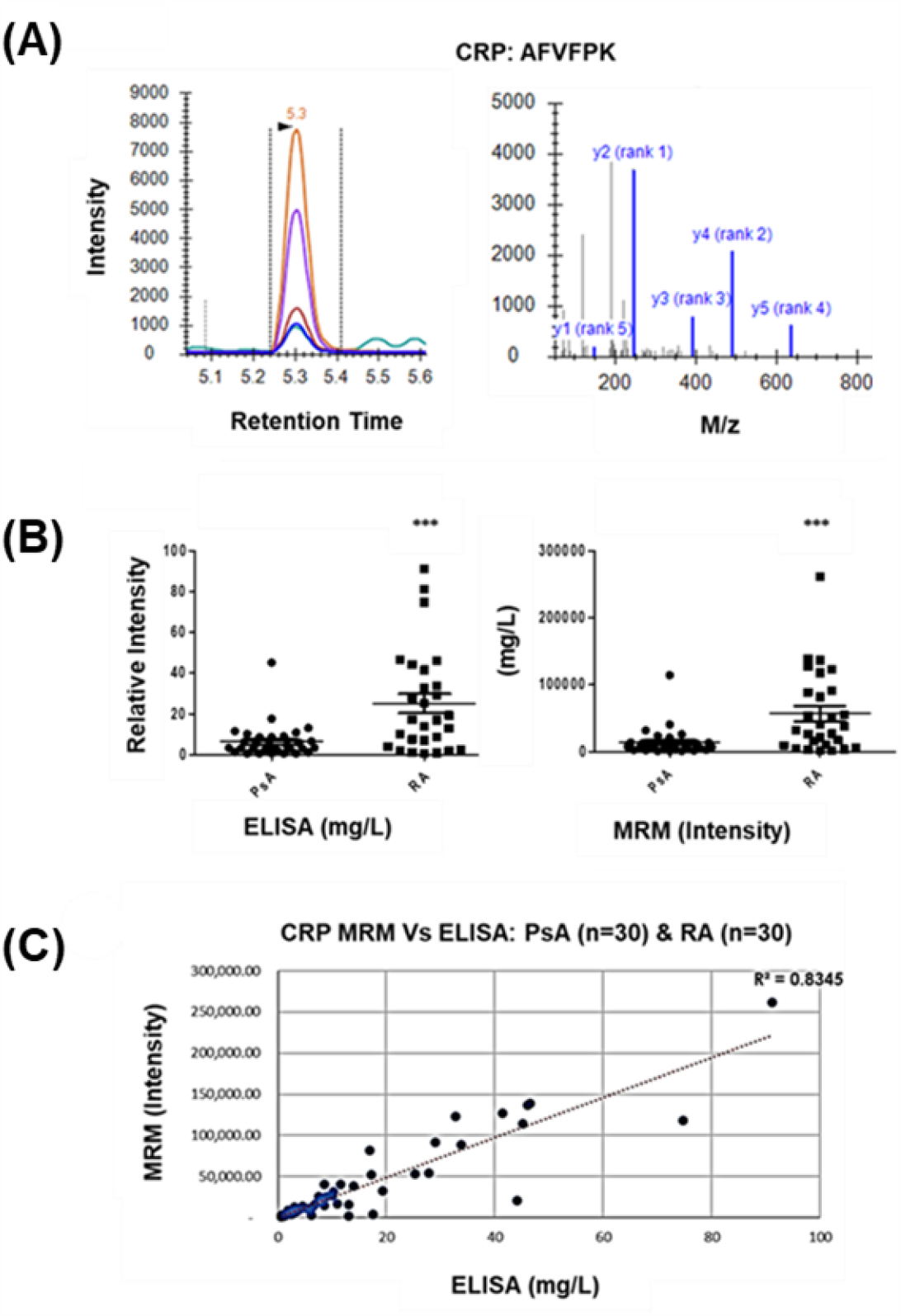
CRP protein expression as measured by MRM and by ELISA. (A) MRM and MS/MS spectrum for CRP, (**B**) Levels of CRP analysed by ELISA (p ≤ 0.009) and MRM (p ≤ 0.006) (**C**) Pearson correlation between ELISA and MRM measurements of CRP (R2 0.8345).

## Discussion

PsA is a complex disease with diverse manifestations; the clinical features observed within individuals with PsA often vary substantially but also overlap with other diseases. While differentiating between PsA and RA can be clinically challenging because of the similarities in their clinical presentation [29], making an accurate diagnosis is increasingly important in order to determine which therapeutic strategy will optimise clinical and radiographic outcomes [30]. Only a few studies have investigated whether there are biomarkers which discriminate between PsA and RA. In one study of synovial tissue, mRNA for VEGF and ANG-2 were elevated in PsA patients compared to RA [31]. Obtaining a synovial biopsy however is an invasive procedure and the discomfort, time and cost associated with tissue sampling makes it undesirable for use in routine clinical practice [31, 32]. More recently, Siebert *et al*. identified 170 urinary peptides which discriminated between patients with long-standing PsA from other arthropathies including early RA with an AUC of 0.97 [33]. These results are promising but urine collection is especially vulnerable to physiological variation arising from diet and liquid intake. Additionally, urine tends to be a very dilute matrix high in salt and low in protein concentration. Thus, in the absence of stepwise workflows for sample concentration and clean up, the quantification of proteins in urine can prove difficult as a result interfering signals present in the matrix [34].

Serum is well recognised as a suitable matrix for biomarker discovery, not least because proteins are shed from relevant affected tissue into the circulation but also because it is readily obtained under standardised operating procedures [35]. Hence, our studies were using serum samples analysed on three proteomic platforms (nLC-MS/MS, SOMAscan and Luminex), each capable of measuring a limited but complementary range of proteins present at different abundance levels. This approach was adopted in order to maximise coverage of serum proteome and to date it is the most comprehensive analysis of the serum proteome in patients with PsA and RA. Although 3 platforms were used to identify putative biomarkers, results from the nLC-MS/MS analysis were the most discriminatory compared to the Luminex and SOMAscan platforms. A potential reason for this is that LC-MS/MS analysis allows for unbiased discovery of biomarkers whereas the other approaches are limited by having fixed panels of protein markers. Furthermore, the SOMAscan platform uses a single aptamer to capture proteins thus potentially reducing the specificity of readouts [36]. It is also possible that the smaller number of patient samples used in the SOMAscan experiments may have constrained the statistical power of the analysis. With respect to the Luminex platform, the 48 carefully selected proteins we measured may not have included key candidate cytokines and chemokines which could be support the discrimination between PsA and RA. The proteins were selected based on their known importance in the pathogenesis of PsA and RA but the panel was limited by the availability of proteins measurable with the in-house assay. With no compelling evidence to justify the time and the cost required to develop further multiplex antibody and/or aptamer assays, we instead focused on the nLC-MS/MS data and performed follow up studies using multiple reaction monitoring (MRM).

MRM represents an excellent tool for supporting large scale multi-protein biomarker studies. It is typically used to refine an initial list of candidate proteins derived from discovery experiments to the subset that may truly address the clinical question under study [37]. MRM analysis is performed on triple quadrupole (QqQ) mass spectrometers which inherently have higher sensitivity and greater linear dynamic range than the orbitrap mass spectrometer used in the discovery experiments here. This boost in sensitivity facilitates the detection of lower abundant proteins in complex samples and therefore reduces the need for sample pre-enrichment steps [38]. Thus, MRM supports more robust workflows as well as time and cost-effective assay development compared to traditional antibody-based approaches. MRM is frequently less sensitive than an equivalent immuno-assay and it was also for this reason that we did not initially attempt to develop MRM assays for putative markers identified only by the SOMAscan or the Luminex analysis [17, 39]. The development of immunoMRM assays to these candidate biomarker proteins represents an obvious way in which improving the performance of the existing panel could be explored [40].

From the two phase of MRM analysis carried described here it was especially interesting to note that a subpanel of 8 proteins (Leucine-rich-alpha-2-glycoprotein, alpha-1-antichymotrypsin, haptoglobin, haptoglobin -related protein, rheumatoid factor C6 light chain, alpha-1-acid glycoprotein 1, coagulation factor XI, thrombospondin-1) were identified as highly discriminatory during the initial verification phase were again confirmed as highly discriminatory during the second evaluation phase. Follow up T-test analysis was performed on this set of proteins and it was found that 7 out of 8 proteins were upregulated in RA compared to PsA during both phases of analysis. Whereas thrombospondin-1 was found to be significantly upregulated in PsA vs RA during the second phase, whereas no significant difference was observed in the initial. This discordance may relate to differences in patient numbers included in the 2 phases or it may relate to the differences in the patients included with the initial phase subjects having early onset, treatment-naive disease whereas those included in the second phase had longer standing disease and were receiving therapy. This in part highlights the advantage of maintaining large panels of proteins for on-going evaluation in patient cohorts. Further analysis of this 8 protein subpanel was carried out using a web-based resource “Search Tool for the retrieval of Interacting Genes/Proteins” (STRING) https://string-db.org/cgi/network.pl revealing the biological functions of these 8 markers of interest, see **Suppl Table 4** for a description. It is interesting to note that this panel is enriched for proteins functionally involved in structural remodelling, angiogenesis, homeostasis and transportation. This perhaps is not surprising since PsA and RA are characterised by an increase in bone turnover and dysregulated angiogenesis.[3, 41]. The radiographic features in PsA and RA can be quite different, with bony erosion observed in both conditions but osteoproliferation only seen in PsA [3]. Hence, in the context of this investigation, it was not unanticipated, that markers of structural remodelling contributed to an algorithm discriminating between individuals with PsA and RA.

Here we demonstrated that a major advantage of using MRM is that it allows the investigator to rapidly adapt a panel to include new candidate biomarkers. We have also shown with our CRP assay developed on MRM over a few days that values highly correlated with those made by ELISA.

Our study has several strengths, including the comprehensive and logical approach to biomarker development. Limitations include the modest number of patient samples in both study phases as well as the absence of healthy and disease controls. Differentiating between PsA from RA is the focus of the current study but it is not the only challenge faced by clinicians, as it can also be challenging to distinguish PsA from other arthropathies and from patients who have skin psoriasis only [14]. This certainly represents a future objective and assessing this biomarker panel in the appropriate additional cohorts is a critical next step. It is noteworthy that the independent cohort included in the second phase of evaluation included patients that had long-standing disease compared to the discovery cohort which were defined as early onset. The performance of the panels may reflect a genuine difference in the protein profile between PsA and RA patients at different stages of disease progression and further work in a larger number of patient samples, will be required to establish if the biomarkers identified here could also be used to distinguish PsA from other diseases and from healthy individuals, at early stages of the disease or as we anticipate, the panel while suited for initial intended use will benefit from further development. Finally, although disease controls weren’t included in our present analysis it is worth highlighting research by Chandran *et al*. that identified differences in serum proteins is patients with PsA compared to patients with osteoarthritis [42] and psoriasis [43]. The protein markers identified in these studies are prime candidates which should be included in future generations of the panel MRM assays.

At present there is no diagnostic test for PsA and as a result, the diagnosis is often late or missed resulting in functional consequences to the patient [12, 44]. With at least 20 % of individuals referred to early arthritis clinics having PsA, there is an urgent need to develop a test to support early detection of this disease [45]. The work described here-in represents a significant contribution towards the development of such a test. Fundamental next steps have been outlined and the MRM approach is ideally suited to support the large-scale studies required to develop and validate a robust panel of discriminatory biomarkers. We believe with further development it will be possible to establish a diagnostic test for PsA which will reduce diagnostic delay, inform treatment selection and improve both short-term and long-term outcomes.

## Data Availability

Data is included in suppl material. Any additional information is available upon request.

## Supplementry Figures & Tables

**Suppl Table 1.**
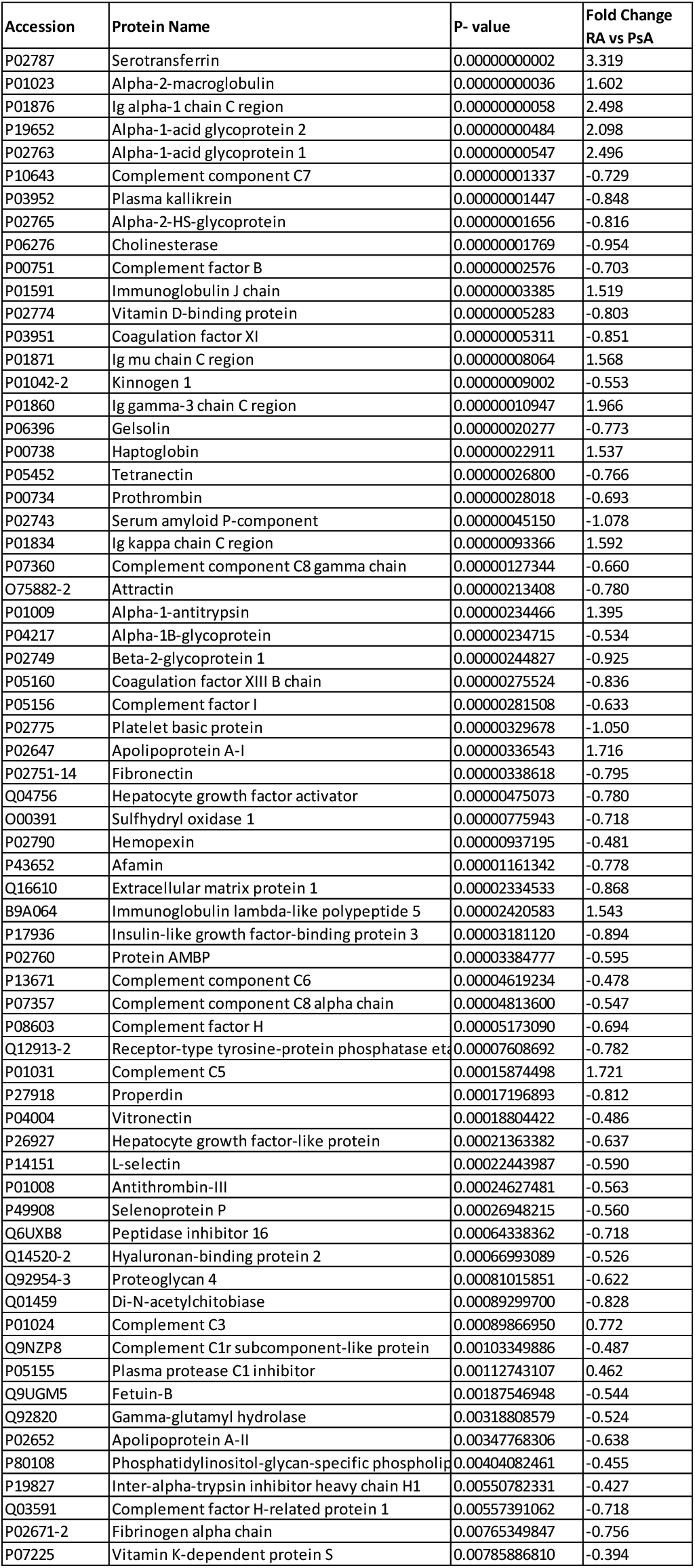
66 proteins were significantly differentially expressed between PsA and RA patients. This table reports a list of 66 proteins that were found to discriminate between patients with PsA (n=30) from those with RA (n=30) during nLC-MS/MS analysis of individual patient serum samples (FDR 0.01, p < 0.01).

**Suppl Table 2.**
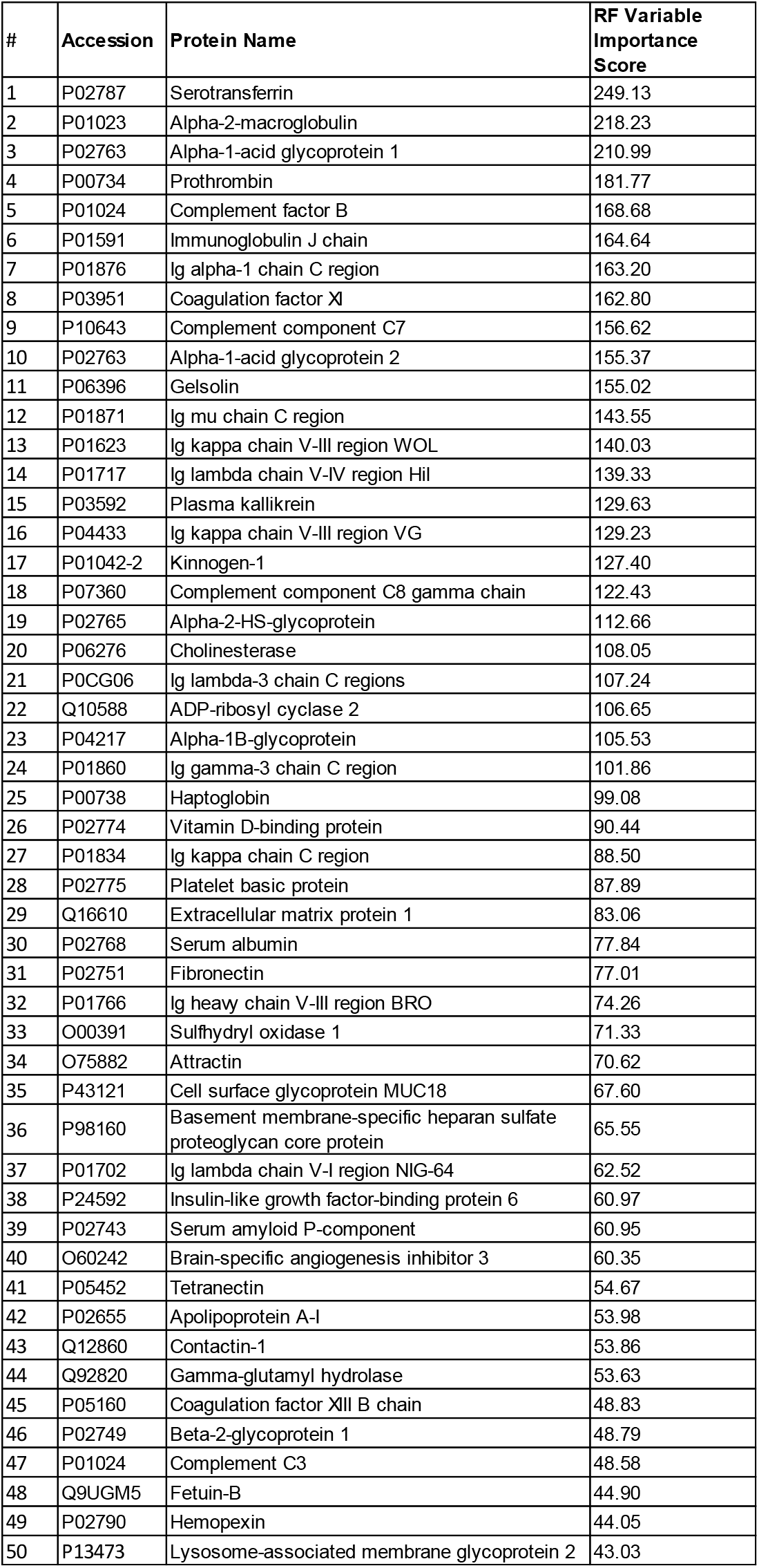
Top 50 ‘nLC-MS/MS’ proteins that discriminate PsA patients from those with RA. Multivariate analysis (RF model) of nLC-MS/MS data revealed a list of serum proteins that could be used to discriminate between patients with PsA and RA (AUC 0.94). The top 50 most discriminatory proteins are reported here. Proteins are listed based on their contribution to the AUC (from high to low).

**Suppl Table 3.** Top 175 ‘SOMAscan’ proteins were significantly differentially expressed between PsA and RA patients. This table reports a list of 175 proteins that were found to discriminate between patients with PsA (n=18) from those with RA (n=18) during SOMAscan analysis of individual patient serum samples (p < 0.05).

| Protein Name | P value (≤) | Fold Change RA vs PsA |
| --- | --- | --- |
| C-X-C motif chemokine 13 | 0.00005 | 2.151346184 |
| Matrix metalloproteinase-8 | 0.00005 | 1.352206659 |
| Apolipoprotein E2 | 0.0001 | 0.823589432 |
| Apolipoprotein E4 | 0.0001 | 0.711015903 |
| Leukocyte immunoglobulin-like receptor subfamily B member 2 | 0.0001 | 1.354612644 |
| C-X-C motif chemokine 10 | 0.0005 | 1.743036335 |
| Elafin | 0.0005 | 0.33417964 |
| Metalloproteinase inhibitor 1 | 0.001 | 1.165906177 |
| Apolipoprotein E | 0.001 | 0.716564956 |
| Apolipoprotein E3 | 0.001 | 0.763093501 |
| Inter alpha trypsin inhibitor heavy chain H4 | 0.001 | 1.142429711 |
| Interleukin 18 | 0.001 | 1.377949845 |
| Granulysin | 0.001 | 1.432742638 |
| Kallikrein 11 | 0.001 | 0.808898431 |
| Fibrinogen | 0.001 | 1.784993976 |
| Angiopoietin 2 | 0.005 | 1.448516326 |
| Granulins | 0.005 | 1.185799393 |
| Tumor necrosis factor receptor 2 | 0.005 | 1.197045355 |
| Tumor necrosis factor $\alpha$ | 0.005 | 1.139440093 |
| Low affinity immunoglobulin gamma Fc region receptor III-B | 0.005 | 1.328470579 |
| Platelet-activating factor acetylhydrolase IB subunit gamma | 0.005 | 0.830888154 |
| Tumor necrosis factor receptor 1 | 0.005 | 1.022389795 |
| Tumor necrosis factor receptor superfamily member 9 | 0.005 | 1.137285699 |
| Phospholipase A2, membrane associated | 0.005 | 1.699232975 |
| Stratifin | 0.005 | 0.886015553 |
| kallikrein 8 | 0.005 | 0.659425376 |
| C-C motif chemokine 18 | 0.005 | 1.615523752 |
| Tumor necrosis factor receptor superfamily member 25 | 0.005 | 1.446772429 |
| Macrophage colony stimulating factor 1 receptor | 0.01 | 1.181312821 |
| Kallistatin | 0.01 | 0.864627283 |
| Acid sphingomyelinase-like phosphodiesterase 3a | 0.01 | 0.890920383 |
| alpha 1 antitrypsin | 0.01 | 1.189217759 |
| Platelet derived growth factor C | 0.01 | 1.237461437 |
| C-C motif chemokine 3 | 0.01 | 1.274622843 |
| Interleukin 5 | 0.01 | 0.888961835 |
| Malate dehydrogenase, cytoplasmic | 0.01 | 0.771971644 |
| Pappalysin-1 | 0.01 | 1.623947009 |
| Insulin-like growth factor binding protein 2 | 0.01 | 1.363747588 |
| Scavenger receptor class F member 1 | 0.01 | 1.198957443 |
| Retinoic acid receptor responder protein 2 | 0.01 | 1.170700539 |
| Fibrinogen gamma chain | 0.01 | 1.500519773 |
| alpha 2-HS-Glycoprotein | 0.01 | 2.174412314 |
| Fibroblast growth factor receptor 1 | 0.01 | 1.043765577 |
| Afamin | 0.01 | 0.848800369 |
| Carbohydrate sulfotransferase 15 | 0.01 | 1.226042089 |
| Protein disulfide isomerase A3 | 0.01 | 0.902420954 |
| GTPase Kras | 0.01 | 1.173925768 |
| Interleukin 22 | 0.01 | 0.875683215 |
| Apoptosis regulator Bcl-2 | 0.01 | 0.720081629 |
| CD48 antigen | 0.01 | 1.10893576 |
| Interleukin 23 | 0.01 | 1.114651543 |
| Growth/differentiation factor 11 | 0.01 | 0.864037986 |
| Peroxiredoxin-6 | 0.01 | 0.725563145 |
| High mobility group protein B 1 | 0.01 | 0.90570097 |
| Dual specificity tyrosine phosphoryl regulated kinase 3 | 0.01 | 0.89248342 |
| Mannan binding lectin serine protease 1 | 0.01 | 0.893863988 |
| Serine/threonine-protein kinase 17B | 0.01 | 0.912919261 |
| Breast cancer anti-estrogen resistance protein 3 | 0.01 | 0.89597676 |
| Dickkopf-like protein 1 | 0.01 | 0.912295607 |
| Carbonic anhydrase I | 0.01 | 0.659578025 |
| Bone Morphogenetic protein 6 | 0.01 | 0.889482833 |
| Marapsin | 0.01 | 0.91683344 |
| Junctional adhesion molecule B | 0.01 | 0.924846483 |
| Acidic leucine rich nuclear phosphoprotein 32 family member B | 0.01 | 1.26712313 |
| Mitogen activated protein kinase 14 | 0.01 | 1.123248061 |
| Cadherin-6 | 0.01 | 0.874389174 |
| Matrilin-3 | 0.01 | 0.904459293 |
| Opioid-binding protein/cell adhesion molecule | 0.01 | 0.917905401 |
| von Willebrand factor | 0.01 | 1.437622207 |
| Prekallikrein | 0.01 | 0.878067125 |
| Calcineurin B a | 0.01 | 0.921660992 |
| Mitogen-activated protein kinase kinase kinase 7 | 0.01 | 0.905300854 |
| Prolactin | 0.01 | 0.902003171 |
| Complement C5 | 0.01 | 1.121719038 |
| Human Dimer D | 0.01 | 1.313578868 |
| Telomeric repeat binding factor 2 interacti protein 1 | 0.01 | 0.816557411 |
| Proteasome subunit alpha | 0.01 | 0.919555921 |
| Stearoyl-CoA desturase | 0.01 | 0.929959093 |
| Tyrosine-protein kinase JAK2 | 0.01 | 1.11275336 |
| Scavenger receptor cysteine-rich type 1 protein M130 | 0.01 | 1.31614313 |
| Follistatin | 0.01 | 0.989096858 |
| Cytidylate kinase | 0.01 | 0.900542524 |
| Renin | 0.01 | 1.438843941 |
| Caherin 15 | 0.01 | 0.898197884 |
| 1,3-beta-glucanosyltransferase gas1 | 0.01 | 0.850860394 |
| Granzyme A | 0.01 | 1.244041525 |
| Kallikrein 14 | 0.05 | 0.919992668 |
| Ferritin | 0.05 | 0.636470764 |
| Mitogen-activated protein kinase 12 | 0.05 | 1.18631665 |
| Apolipoprotein A 1 | 0.05 | 0.8599308 |
| Lymphatic vessel endothelial hyaluronic acid receptor | 0.05 | 1.197013536 |
| Ectonucleoside triphosphate diphosphohydrolase 3 | 0.05 | 0.905849506 |
| WAP, Kazal, immunoglobulin, Kunitz and NTR domain-containing protein 1 | 0.05 | 0.853000848 |
| Superoxide dismutase | 0.05 | 0.81705517 |
| Tumor necrosis factor receptor superfamily member 9 | 0.05 | 1.136152512 |
| kallikrein 13 | 0.05 | 0.903974531 |
| Creatine kinase M type | 0.05 | 0.89797268 |
| Ephrin type-B receptor 4 | 0.05 | 0.925039795 |
| Choriogonadotropin subunit beta 3 | 0.05 | 0.888157763 |
| Choriogonadotropin subunit beta 3 | 0.05 | 4.408355687 |
| Neurexophilin-1 | 0.05 | 1.137754083 |
| Kremen protein 2 | 0.05 | 0.82612986 |
| Basigin | 0.05 | 0.914290822 |
| Abelson tyrosine-protein kinase 2 | 0.05 | 0.90155234 |
| IgD | 0.05 | 1.679004239 |
| Prothrombin | 0.05 | 0.770698711 |
| Importin subunit alpha 1 | 0.05 | 1.098775371 |
| Fibroblast growth factor 16 | 0.05 | 0.767549544 |
| Hemoglobin | 0.05 | 0.705353445 |
| Non-receptor tyrosine kinase | 0.05 | 0.919807205 |
| Pituitary adenylate cyclase-activating polypeptide | 0.05 | 0.898662293 |
| C-C chemokine 23 | 0.05 | 1.237132858 |
| Fibroblast growth factor 8 | 0.05 | 0.723007869 |
| Interlukin 6 | 0.05 | 1.310845892 |
| C-C motif chemokine 23 | 0.05 | 1.185653759 |
| Delta-like-protein 1 | 0.05 | 1.066152382 |
| C34 gp41 HIV Fragment | 0.05 | 1.114347166 |
| Collectin-12 | 0.05 | 0.9135605 |
| Stabilin 2 | 0.05 | 0.915417588 |
| Ectonucleoside triphosphate diphosphohydrolase 1 | 0.05 | 0.941522964 |
| Apolipoprotein D | 0.05 | 0.930863567 |
| Receptor-type tyrosine protein kinase 3 | 0.05 | 0.897868651 |
| Alpha-(1,3)-fucosyltransferase 5 | 0.05 | 1.303568345 |
| Histone acetyltransferase KAT6A | 0.05 | 0.930994218 |
| kinnogen 1 | 0.05 | 0.878275356 |
| Tryptase gamma | 0.05 | 0.870826658 |
| Interleukin18 binding protein | 0.05 | 1.219927556 |
| Teratocarcinoma-derived growth factor 1 | 0.05 | 0.873444058 |
| cGMP-inhibited 3',5'-cyclic phosphodiesterase A | 0.05 | 0.905336717 |
| Fibroblast growth factor 23 | 0.05 | 1.175393989 |
| Cadherin-12 | 0.05 | 0.818190025 |
| Persulfide dioxygenase ETHE1, mitochondrial | 0.05 | 1.080652246 |
| Discoidin domain receptor 2 | 0.05 | 0.932402477 |
| Serine/threonine-protein kinase MRCK beta | 0.05 | 0.895874112 |
| Persephin | 0.05 | 0.915744525 |
| cGMP-stimulated PDE | 0.05 | 0.928438289 |
| Peptidyl-prolyl cis-trans isomerase B | 0.05 | 0.909410267 |
| Leucine-rich repeat transmembrane protein FLRT1 | 0.05 | 0.874881829 |
| Angiopoietin-1 receptor | 0.05 | 1.133466793 |
| Tumor necrosis factor ligand superfamily member 13B | 0.05 | 1.318831839 |
| Bone morphogenetic protein 1 | 0.05 | 0.861501004 |
| A disintegrin and metalloproteinase with thrombospondin motifs 15 | 0.05 | 0.923496633 |
| Collagen alpha-1(XVIII) chain | 0.05 | 1.116382129 |
| Interleukin 6 receptor | 0.05 | 1.161712342 |
| Thymic stromal lymphopoietin | 0.05 | 1.701846315 |
| Vascular endothelial growth factor A | 0.05 | 1.307321244 |
| Hepatocyte growth factor receptor | 0.05 | 1.115242093 |
| Proteasome subunit alpha type-1 | 0.05 | 0.949679601 |
| Nucleolar pre-ribosomal-associated protein 1 | 0.05 | 1.20486678 |
| Lymphocyte activation gene 3 protein | 0.05 | 1.298871612 |
| Sialic acid-binding Ig-like lectin 14 | 0.05 | 1.254231462 |
| Prosaposin | 0.05 | 0.905866435 |
| Proteasome subunit alpha type-6 | 0.05 | 0.824069519 |
| Cathepsin S | 0.05 | 1.133412457 |
| Histone-lysine N-methyltransferase EHMT2 | 0.05 | 0.89539073 |
| Ephrin-A4 | 0.05 | 1.118455027 |
| Coagulation Factor X | 0.05 | 0.909135559 |
| OCIA domain-containing protein 1 | 0.05 | 0.906278293 |
| Estrogen receptor | 0.05 | 0.76749157 |
| Intercellular adhesion molecule 3 | 0.05 | 0.91017701 |
| beta 2 microglobulin | 0.05 | 1.152800158 |
| Carbonic anhydrase 6 | 0.05 | 0.702195224 |
| Tumor necrosis factor receptor superfamily member 19 | 0.05 | 1.238543031 |
| Protein 4.1 | 0.05 | 0.619582347 |
| Disintegrin and metalloproteinase domain-containing protein 12 | 0.05 | 1.169252858 |
| Ubiquitin-conjugating enzyme E2 L3 | 0.05 | 0.802367055 |
| Leukocyte immunoglobulin-like receptor subfamily B member 1 | 0.05 | 1.218604813 |
| Cytokine receptor common subunit gamma | 0.05 | 0.749869438 |
| Estradiol 17-beta-dehydrogenase 1 | 0.05 | 0.904544796 |
| Lysozyme | 0.05 | 1.167264513 |
| Cell adhesion molecule 1 | 0.05 | 0.910728088 |
| T-lymphocyte activation antigen CD86 | 0.05 | 1.252274616 |
| C-X-C motif chemokine 11 | 0.05 | 1.348865796 |
| C-type lectin domain family 7 member<br>A | 0.05 | 0.913065021 |
| CD209 antigen | 0.05 | 1.155602828 |

**Suppl Figure 1:**
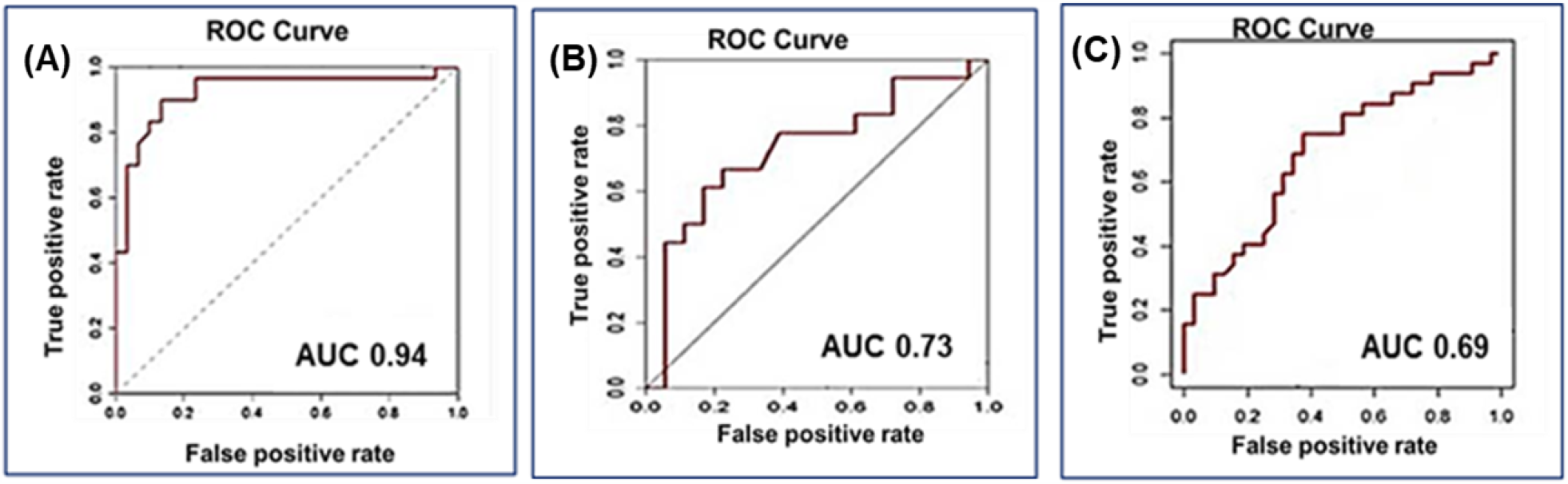
ROC analysis of (A) LC-MS/MS (n=60) (B) SOMAscan (n=36), (C) Luminex (n=64).

**Suppl Fig 2.**
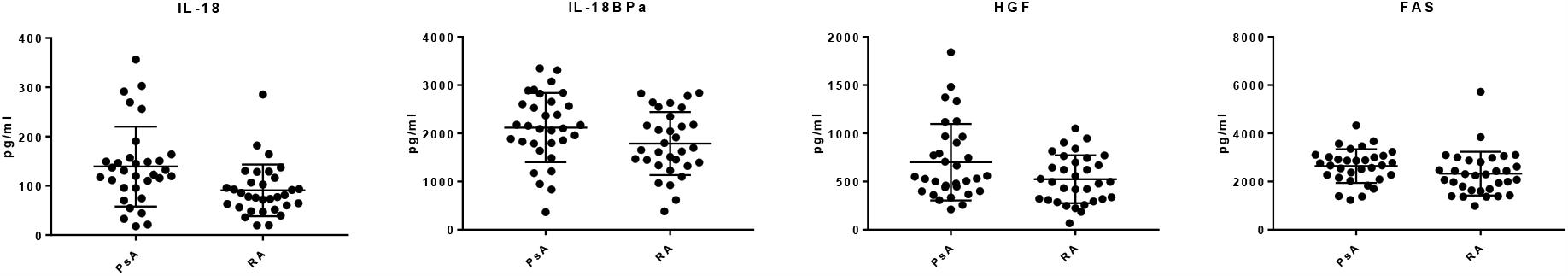
Serum proteins measured by Luminex analysis which were significantly differentially expressed between PsA and RA patients. Luminex analysis of serum samples revealed (A) IL-18 (p ≤ 0.001), Il-18 BPa, HGF and FAS (p ≤ 0.05) were differentially expressed between PsA (n=32) and RA(n=32).

**Suppl Figure 3.**
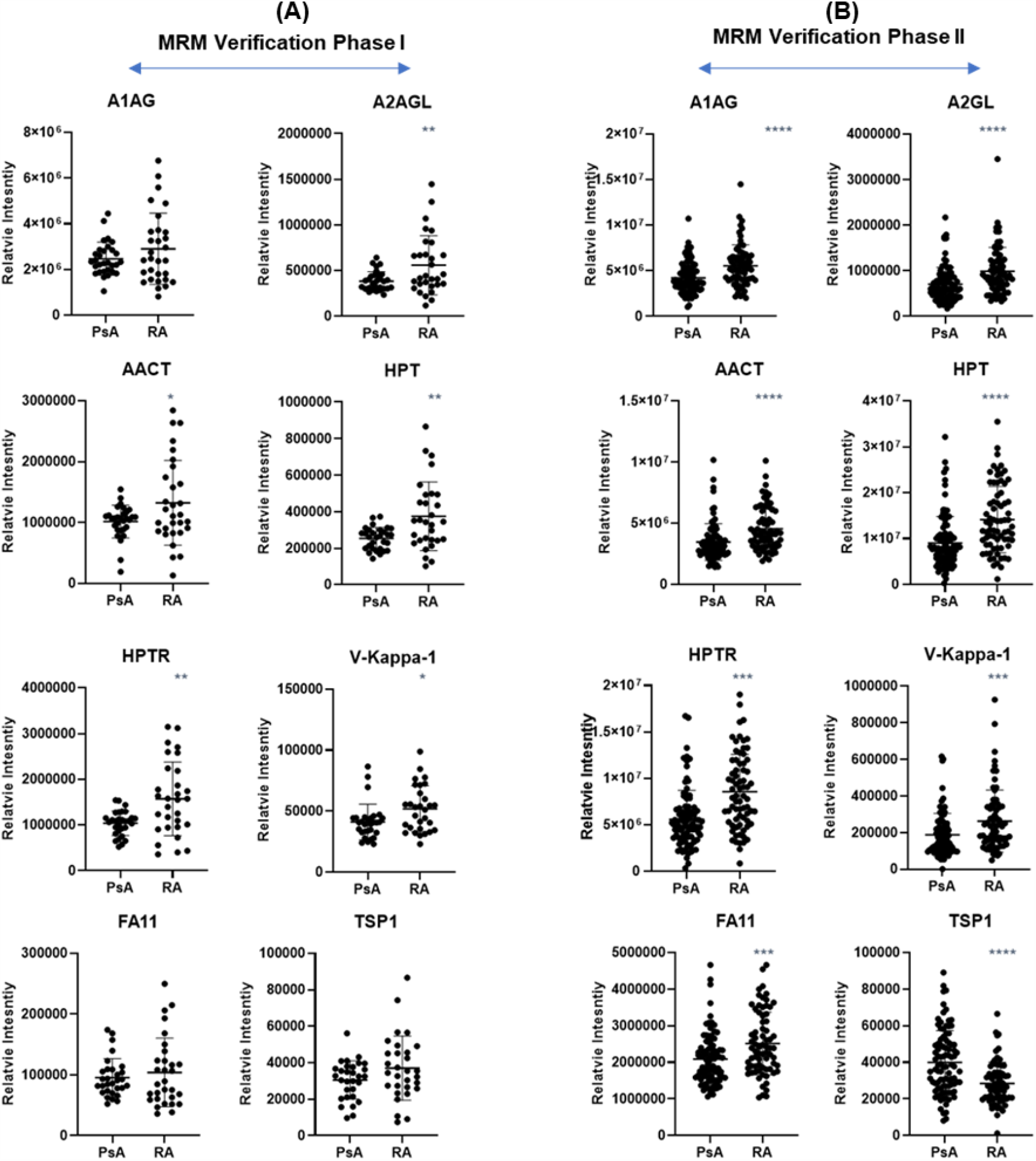
Pattern of expression changes in proteins measured by MRM. Where two peptides were available per protein, plots were generated based of the summed peptide intensity. **(A)** During MRM phase I A1AG, FA11 and TSP1 were not significantly differently expressed between PsA and RA patients. Proteins; A2AGL (p < 0.006), AACT (p < 0.020), HPT (p < 0.001), HPTR (p <0.001), V-Kappa-1 (p < 0.015) were significantly upregulated in RA. **(B)** During MRM Phase II proteins; A1AG (p < 0.00001), A2AGL (p < 0.00001), AACT (p < 0.00001), HPT (p < 0.0001), HPTR (p < 0.00001), V-Kappa-1 (p < 0.0001) and FA11 were significantly upregulated in RA while TSP1 was significantly upregulated in PsA (p < 0.00001).

**Suppl Table 4.** Functional roles of discriminatory proteins included in MRM analysis. Table was generated based of results from STRING software.

| Protein | Biological Role | General Function |
| --- | --- | --- |
| A1AG | functions as transport protein in the blood stream. Binds various ligands in the interior of its beta-barrel domain. Also binds synthetic drugs and influences their distribution and availability in the body. Appears to function in modulating the activity of the immune system during the acute-phase reaction | Transport/homeostasis |
| A2AGL | Promotes endocytosis, possesses opsonic properties and influences the mineral phase of bone. Shows affinity for calcium and barium ions. | Structural Remodeling |
| AACT | Is a serpin peptidase inhibitor it can inhibit neutrophil cathepsin G and mast cell chymase, both of which can convert angiotensin-1 to the active angiotensin-2. | Structural Remodeling |
| FA11 | Triggers the middle phase of the intrinsic pathway of blood coagulation by activating factor IX. | Structural Remodeling |
| HPT | As a result of hemolysis, hemoglobin is found to accumulate in the kidney and is secreted in the urine. Haptoglobin captures, and combines with free plasma hemoglobin to allow hepatic recycling of heme iron and to prevent kidney damage. Haptoglobin also acts as an Antimicrobial; Antioxidant, has antibacterial activity and plays a role in modulating many aspects of the acute phase response. Hemoglobin/haptoglobin complexes are rapidly cleared by the macrophage CD163 scavenger receptor expressed on the surface of liver Kupfer cells through an endocytic lysosomal degradation | Transport/homeostasis |
| HPTR | The function of HPTR overlaps with HPT however HPTR has an affinity for high density lipoprotein and thus contributes to the clearance of cholesterol from blood | Transport/homeostasis |
| TSP1 | Adhesive glycoprotein that mediates cell-to-cell and cell-to-matrix interactions. Binds heparin. May play a role in dentinogenesis and/or maintenance of dentin and dental pulp (By similarity). Ligand for CD36 mediating antiangiogenic properties. Plays a role in ER stress response, via its interaction with the activating transcription factor 6 alpha (ATF6) which produces adaptive ER stress response factors | Structural Remodeling |
| V-Kappa-1 | Promotes complement fixation and thus endocytosis. | Structural Remodeling |

## Suppl Doc 1

### Standard Operating Procedure for Serum Processing from Whole Blood

This Standard Operating Procedure is developed in accordance with the International Conference on Harmonisation (ICH) Harmonised Tripartite guideline for Good Clinical Practice (GCP) and Clinical Trials on Products for Human Medicinal Use Regulations 2004 to 2006 (SI 190 of 2004 & SI 374 of 2006), where applicable.

The Standard Operating Procedure applies to all staff involved in MIAMI projects.

**Objectives:** To ensure the biomarker discovery and validation (mRNA, miRNA and proteomic) samples are collected, processed and stored in an appropriate manner.

**Scope:** This SOP applies to the proteomic studies of MIAMI (WP4)

**Definition:** Collection, processing and storage of study samples.

**Prepared by:** Stephen Pennington, Conway Institute, University College Dublin with input from Phil Gallagher, St Vincent’s University Hospital, Dublin

**Procedure:**

**All sample processing information should be recorded and stored in the sample database associated with the sample i**.**d**.**/labels**.

1. Gloves and laboratory coat should be worn throughout sample processing to protect the sample from contamination (this includes labelling the tubes prior to sample processing). All tubes and pipettes should be covered at all times to minimise dust contamination.
2. Blood samples should be collected using a vacutainer to avoid haemolysis. Three 7.5 ml / 10 ml anti-coagulant free tube (red top serum tubes) will be used to collect serum.
3. Filled blood tubes should be left to sit upright after blood is drawn at room temperature for a minimum of 30 to a maximum of 60 minutes to allow the clot to form.
4. Centrifuge the blood sample at the end of the clotting time (30-60 minutes) for 15 minutes at 1800 g at room temperature. If the blood is not centrifuged immediately after the clotting time, the tubes should be refrigerated (4°C) for no longer than 4 hours prior to centrifugation. In all cases the time of clotting and temperature the samples were subjected to should be recorded.
5. Using pipette transfer the serum from the vacutainers into a larger tube and mix. Pipette serum into the labelled cryovials in aliquots of 1000 μl. Close the cap on the vials tightly. This process should be completed within 1 hour of centrifugation. Note: To avoid picking up red blood cells when aliquoting keep the pipet above the red blood cell layer and leaving a small amount of serum in the tube.
6. Samples should be labelled as described in MIAMI SOP:OO2.
7. Place all aliquots upright in a specimen box and store at -80°C.

